# Development of a rapid SARS-CoV-2 neutralisation test by detecting antibodies that block interaction of spike protein receptor-binding domain (RBD) with angiotensin-converting enzyme 2 (ACE2)

**DOI:** 10.1101/2021.06.03.21255116

**Authors:** Ciaran Richardson, Sarah Gildea, Stephen Harkin, Annie Gallagher, Eimear Conroy, Lorraine McClafferty, Eibhlin McCole, El Ouard Benchikh, Cherith N. Reid, Marshall Dunlop, Sharon Savage, Philip Lowry, Jim Curry, Robert I. McConnell, John V. Lamont, Stephen P. FitzGerald

## Abstract

The urgent need for a rapid and reliable Severe Acute Respiratory Syndrome Coronavirus-2 (SARS-CoV-2) neutralising antibody detection test compatible with routine clinical laboratory testing currently exists. This is necessary to provide accurate estimates of immunity and to monitor vaccine effectiveness. Utilising Biochip Array Technology (BAT), the Randox SARS-CoV-2 Biochip proxy virus neutralisation test (pVNT) has been developed. Immobilising SARS-CoV-2 Spike RBD on the Biochip surface, innovative assay design enables direct sample addition to the Biochip well without the need for off-board sample pre-incubation step. Results are reported within 1.5 hours and testing is carried out without the prerequisite of live virus or biosafety level 3 (BSL3) laboratory facilities. In this study, assay validation is performed using recombinant antibodies and clinical samples and an excellent correlation against conventional virus neutralisation methods is established (100% clinical specificity and 98% clinical sensitivity). Serial dilution of samples with high neutralising antibody levels demonstrate end-point sample dilutions comparable with those obtained using the SARS-CoV-2 microneutralisation test. Species independent neutralising antibody detection capacity of the SARS-CoV-2 Biochip pVNT is also demonstrated. The findings of this study exemplify the utility of the SARS-CoV-2 Biochip pVNT as a robust and reliable method for the accurate measurement of neutralising antibodies against SARS-CoV-2 and the availability of this test can now positively impact current testing deficiencies in this area.

## Introduction

Coronavirus Disease 2019 (COVID-19) which is caused by SARS-CoV-2 infection was first recognized in December 2019 in Wuhan, China^1^. On 30^th^ January 2020, The World Health Organization (WHO) stated that COVID-19 constituted a “public health emergency of international concern”, latterly, on 11^th^ March, it was declared a pandemic. Infection is initiated when the viral trimeric Spike (S) protein binds to ACE2 subsequently, proteolytic cleavage of the S protein by TMPRSS2 or cathepsin L mediates viral entry^2^. Molecular detection using polymerase chain reaction (PCR) continues to play a pivotal role enabling “gold standard” diagnostic testing while next-generation sequencing is being increasingly deployed to monitor genetic variants that are arising globally^3^. The recently emergent variants; UK, B.1.1.7; South Africa, B.1.351; and Brazil, P.1. exhibit multiple mutations in the immunodominant Spike protein RBD and these constitute significant concern for their potential to induce immune evasion^4^. This has significant implications for the ongoing global effort to produce effective vaccines and the concept of establishing societal immunity^5^.

The gold standard virus neutralisation test (VNT) which detect neutralising antibodies in a subject’s blood sample must be completed in a specialized BSL3 containment laboratory because it uses infectious SARS-CoV-2 virus with live cell invasion; the test format is laborious taking 2-4 days to complete^6^. Deployment of pseudovirus-based virus neutralisation tests which can be completed with biosafety level 2 (BSL2) containment are equally tedious and time consuming^7^. Consequently, there is an urgent need to develop a rapid, reliable serological based SARS-CoV-2 neutralising antibody detection immunoassay.

In this study, we report the development of a rapid proxy virus neutralisation test (pVNT) using Randox’s proprietary BAT, which can detect SARS-CoV-2 neutralising antibodies in a subject’s blood sample compatible with routine clinical laboratory testing. The *in vitro* pVNT concept is based on the *in vivo* interaction of spike protein RBD with ACE2; this highly specific protein-protein interaction will be blocked if a subject has developed an antibody repertoire that prevents the interaction between spike protein RBD and ACE2 i.e. neutralising antibodies. Our rapid pVNT format mimics that of the classic VNT with results generated within 1.5 hours.

## Materials and Methods

### Human and animal samples

Seronegative human serum and plasma samples were collected from individuals pre COVID-19 (before October 2019). Seropositive human serum and plasma samples were collected from SARS-CoV-2 patients at fixed time points following RT-PCR confirmation of SARS-CoV-2 infection. Human serum and plasma samples containing antibodies against endemic human coronaviruses (HKU1, OC43, 229E, NL63) and other non-coronavirus related pathogens were also collected.

Sheep antisera was collected following immunisation with SARS-CoV-2 Spike RBD (GenScript, Z03483) and SARS-CoV-2 Nucleocapsid protein (GenScript, Z03488). Samples were collected on the day of immunisation followed by 3, 6, 10, 13, 20, 28 and 52 days post immunisation. Immunisation and the collection of samples from sheep was carried out under Animals (Scientific Procedures) Act 1986 – Project License number PPL2797.

### Recombinant antibodies

Humanised IgG recombinant antibodies SARS-CoV-1 RBD Antibody 1 (RAB0001), SARS-CoV-1 RBD Antibody 2 (RAB0003), SARS-CoV-2 RBD Antibody 1 (RAB0006) and SARS-CoV-2 RBD Antibody 2 (RAB0005) were supplied by Randox Laboratories Ltd.

### Sample characterisation using SARS-CoV-2 microneutralisation assay

Two-fold serial dilution of sample was mixed 1:1 with SARS-CoV-2 virus (Germany/BavPat1/2020) and pre-incubated at 37°C for 1 hour to allow neutralisation. The virus-sample mixture was transferred into Vero-E6 cells monolayers and incubated at 37°C for 1 hour. One-hour post-infection, the inoculum was replenished with fresh growth medium and plates were incubated overnight after which the Vero-E6 cells were fixed and immunostained to visualise infected cells. MN80 titres were calculated according to the method described by Zielinski et al^8^ and based on three sample replicates.

### Sample characterisation using SARS-CoV-2 IgG (RBD & NP) assay

SARS-Cov-2 RBD IgG antibody levels were measured using the SARS-CoV-2 IgG (RBD & NP) array kit (Randox Laboratories Ltd, EV4447) as per the manufacturer’s instructions. Samples with a % positivity (SP%) > 50% are reported as seropositive for SARS-CoV-2 RBD IgG antibodies.

### SARS-CoV-2 Biochip pVNT (RBD immobilised on Biochip)

Virus neutralisation antibodies were detected in serum and plasma samples using a blocking chemiluminescent Biochip-based immunoassay on the Evidence Investigator Biochip analyser (SARS-CoV-2 Surrogate Virus Neutralising Assay EV4454, Evidence Investigator analyser EV3602, Randox Laboratories Ltd). The core technology is an established solid-state device comprising a carrier holding nine Biochips, each containing immobilised RBD antigen located at a discrete test region (DTR). SARS-CoV-2 neutralising antibodies if present, will bind RBD, inhibiting its ability to interact with ACE2 labelled horseradish peroxidase (ACE2 HRP). Light signal generated from the RBD immobilised DTR is detected using digital imaging technology as previously described^9^ and is inversely proportional to the amount of SARS-CoV-2 neutralising antibody present in a sample.

Briefly, 288μL assay diluent and 12μL sample or positive/negative control (1/25 in Biochip well sample dilution) were applied directly to the Biochip. After incubation at 37 °C and 370 rpm for 30 min on a thermoshaker (EV702-527, Randox Laboratories Ltd), the supernatant was discarded. Following two quick and four 2-min washing cycles, 300μL of ACE2 HRP was added to each well and incubated at 37 °C and 370 rpm for 30 min. After a second wash cycle (same as above), 250μL signal reagent (1:1 mixture of peroxide/ luminol solution) was added to each Biochip well and incubated for 2 min in the dark. Emitted light from the RBD antigen immobilised DTR was detected by a charged coupled device (CCD) camera and signal was quantified using the Evidence Investigator System and its imaging processing software. Percentage inhibition (% inhibition) was calculated for each sample relative to the negative control. Samples with % inhibition > 50% are reported as seropositive for SARS-CoV-2 neutralising antibodies.

### SARS-CoV-2 Biochip pVNT (ACE2 immobilised on Biochip)

In the present study, an assay format utilising ACE2 antigen (Randox Laboratories Ltd, RCP9642) immobilised on the Biochip was also explored. Briefly, sample or positive/negative control was incubated off-board at a ratio of 16μL to 384μL assay diluent containing RBD (Randox Laboratories Ltd, RCP9639) labelled with HRP for 30 minutes at 37 °C. Following incubation 300μL of this preparation was added to the Biochip well containing immobilised ACE2 and incubated at 37 °C and 370 rpm for 30 min on a thermoshaker. Following two quick and four 2-min washing cycles, 250μL signal reagent (1:1 mixture of peroxide/ luminol solution) was added to each well and incubated for 2 min in the dark. Emitted light from the ACE2 antigen immobilised DTR was detected by a CCD camera, and signal was quantified using the Evidence Investigator System and its imaging processing software. Percentage inhibition (% inhibition) was calculated for each sample relative to the negative control.

### Statistics

The correlation between the SARS-CoV-2 Biochip pVNT and SARS-CoV-2 microneutralisation assay was calculated with Pearson’s correlation coefficients on MedCalc version 19.0.4. using logarithmically SARS-CoV-2 microneutralisation transformed assay data.

## Results

### Demonstration of bidirectional interaction between Spike RBD and ACE2 employing BAT

Employing BAT, we demonstrated functional interaction between ACE2 immobilized on the Biochip surface and HRP labelled RBD (Figure 1A, Table 1A). Furthermore, it was demonstrated that this interaction could be disrupted using various recombinant neutralising antibodies as well as SARS-CoV-2 RBD IgG seropositive samples. Two SARS-CoV-1 RBD antibodies (SARS-CoV-1 RBD Antibody 1, SARS-CoV-1 RBD Antibody 2) exhibited 36.0% and 98.5% inhibition respectively whereas two SARS-CoV-2 RBD antibodies (SARS-CoV-2 RBD Antibody 1, SARS-CoV-2 RBD Antibody 2) utilized at the same fixed concentration demonstrated 100% inhibition of the ACE2/RBD interaction. Clinical samples classified as SARS-CoV-2 RBD IgG seropositive disrupted the ACE2 interaction with % inhibition ranging from 10.2% to 99.9%. The possibility of orientating the assay in the opposite direction was therefore considered (Figure 1B) thereby potentially eliminating the requirement for an offboard pre-incubation step. Functional interaction was successfully demonstrated between RBD immobilized on the Biochip surface and HRP labelled ACE2 with comparable disruption of this interaction achieved employing the same suite of recombinant neutralising antibodies and clinical samples that disrupted the reverse assay orientation (Table 1B). It was decided to proceed with development of a pVNT that exploits the interaction between RBD and ACE2 whereby RBD is immobilized on the Biochip surface as opposed to ACE2.

**Figure 1:**
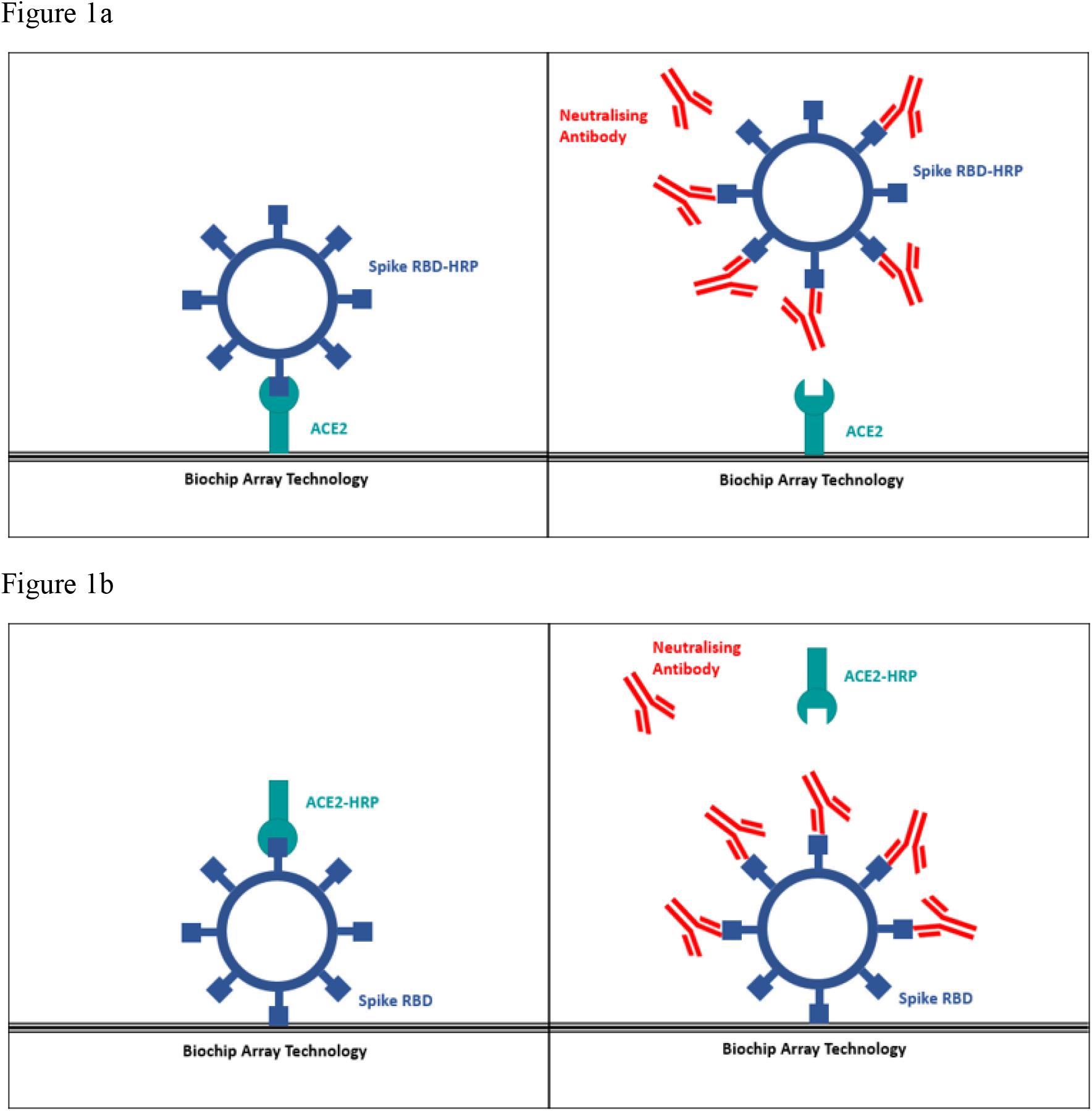
Principle of SARS-CoV-2 Biochip pVNT. 1a: ACE2 immobilised on Biochip. Anti-SARS-CoV-2 neutralising antibodies block SARS-CoV-2 Spike RBD HRP from binding to ACE2 on Biochip. 1b: SARS-CoV-2 Spike RBD immobilised on Biochip. Anti-SARS-CoV-2 neutralising antibodies block ACE2 HRP from binding to SARS-CoV-2 RBD on Biochip.

**Table 1:**
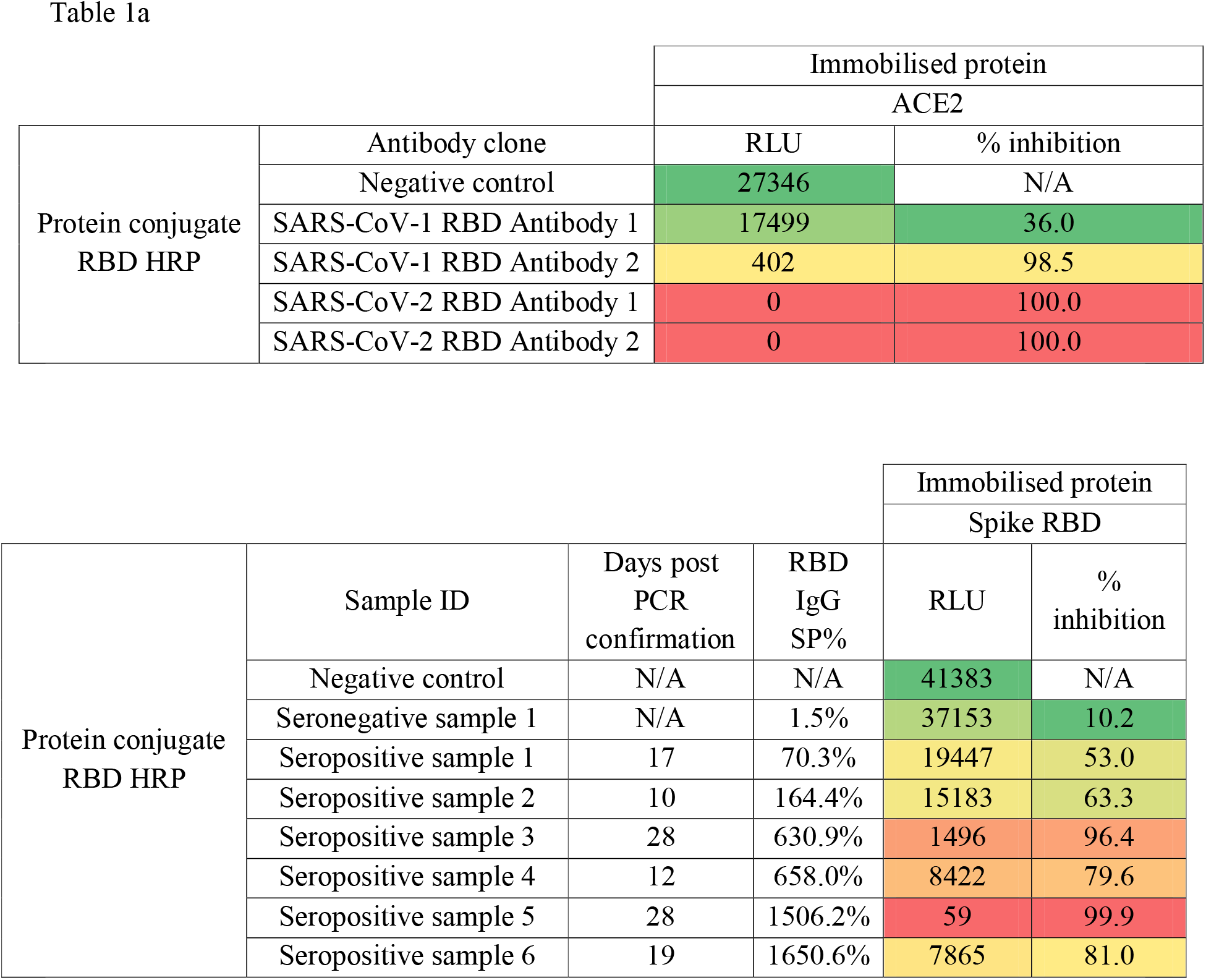
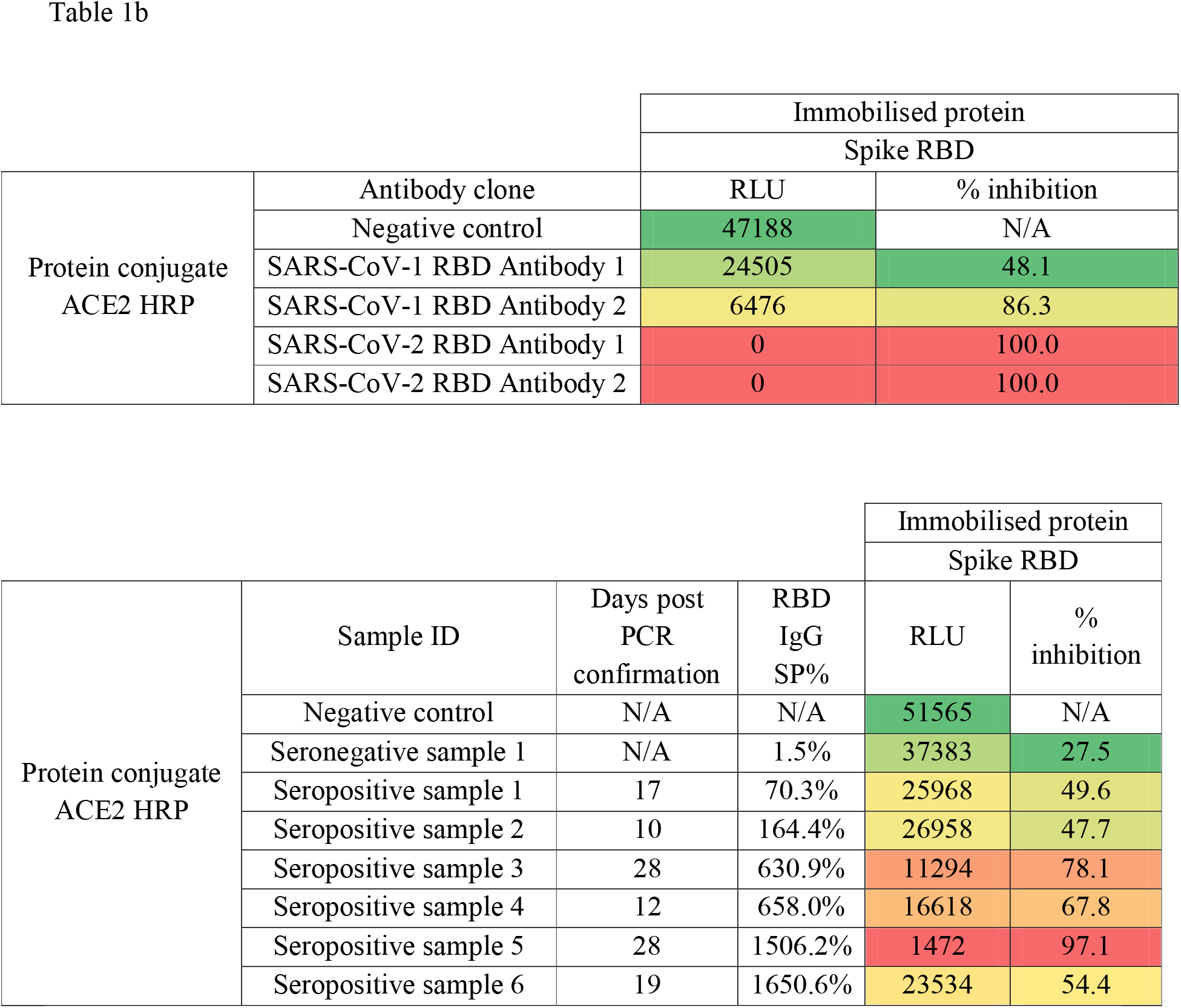
Validation of SARS-CoV-2 Biochip pVNT using SARS-CoV-2 recombinant antibodies and samples collected from COVID-19 patients confirmed positive by RT-PCR. 1a: Result obtained with ACE2 immobilised on Biochip. 1b: Result obtained with SARS-CoV-2 RBD immobilised on Biochip.

|  |  | Immobilised protein |  |
| --- | --- | --- | --- |
|  |  | ACE2 |  |
| Protein conjugate<br>RBD HRP | Antibody clone | RLU | % inhibition |
|  | Negative control | 27346 | N/A |
|  | SARS-CoV-1 RBD Antibody 1 | 17499 | 36.0 |
|  | SARS-CoV-1 RBD Antibody 2 | 402 | 98.5 |
|  | SARS-CoV-2 RBD Antibody 1 | 0 | 100.0 |
|  | SARS-CoV-2 RBD Antibody 2 | 0 | 100.0 |

|  |  |  |  | Immobilised protein |  |
| --- | --- | --- | --- | --- | --- |
|  |  |  |  | Spike RBD |  |
| Protein conjugate<br>RBD HRP | Sample ID | Days post<br>PCR<br>confirmation | RBD<br>IgG<br>SP% | RLU | %<br>inhibition |
|  | Negative control | N/A | N/A | 41383 | N/A |
|  | Seronegative sample 1 | N/A | 1.5% | 37153 | 10.2 |
|  | Seropositive sample 1 | 17 | 70.3% | 19447 | 53.0 |
|  | Seropositive sample 2 | 10 | 164.4% | 15183 | 63.3 |
|  | Seropositive sample 3 | 28 | 630.9% | 1496 | 96.4 |
|  | Seropositive sample 4 | 12 | 658.0% | 8422 | 79.6 |
|  | Seropositive sample 5 | 28 | 1506.2% | 59 | 99.9 |
|  | Seropositive sample 6 | 19 | 1650.6% | 7865 | 81.0 |
|  |  | Spike RBD |  |
| Protein conjugate<br>ACE2 HRP | Antibody clone | RLU | % inhibition |
|  | Negative control | 47188 | N/A |
|  | SARS-CoV-1 RBD Antibody 1 | 24505 | 48.1 |
|  | SARS-CoV-1 RBD Antibody 2 | 6476 | 86.3 |
|  | SARS-CoV-2 RBD Antibody 1 | 0 | 100.0 |
|  | SARS-CoV-2 RBD Antibody 2 | 0 | 100.0 |

|  |  |  |  | Immobilised protein |  |
| --- | --- | --- | --- | --- | --- |
|  |  |  |  | Spike RBD |  |
| Protein conjugate<br>ACE2 HRP | Sample ID | Days post<br>PCR<br>confirmation | RBD<br>IgG<br>SP% | RLU | %<br>inhibition |
|  | Negative control | N/A | N/A | 51565 | N/A |
|  | Seronegative sample 1 | N/A | 1.5% | 37383 | 27.5 |
|  | Seropositive sample 1 | 17 | 70.3% | 25968 | 49.6 |
|  | Seropositive sample 2 | 10 | 164.4% | 26958 | 47.7 |
|  | Seropositive sample 3 | 28 | 630.9% | 11294 | 78.1 |
|  | Seropositive sample 4 | 12 | 658.0% | 16618 | 67.8 |
|  | Seropositive sample 5 | 28 | 1506.2% | 1472 | 97.1 |
|  | Seropositive sample 6 | 19 | 1650.6% | 23534 | 54.4 |

### Stepwise disruption of RBD/ACE2 interaction using two recombinant neutralising antibodies that bind SARS-CoV-2 RBD

To further demonstrate the functionality of the proposed pVNT we successfully demonstrated that two recombinant neutralising antibodies (SARS-CoV-2 RBD Antibody 1 and SARS-CoV-2 RBD Antibody 2) abrogated ACE2 interaction with RBD immobilized on the Biochip surface in a dose dependent manner (Figure 2). Complete inhibition of the RBD/ACE2 interaction was demonstrated at concentrations less than 10 ug/ml for both antibodies. This data further substantiates the functional validity of the proposed assay format.

**Figure 2:**
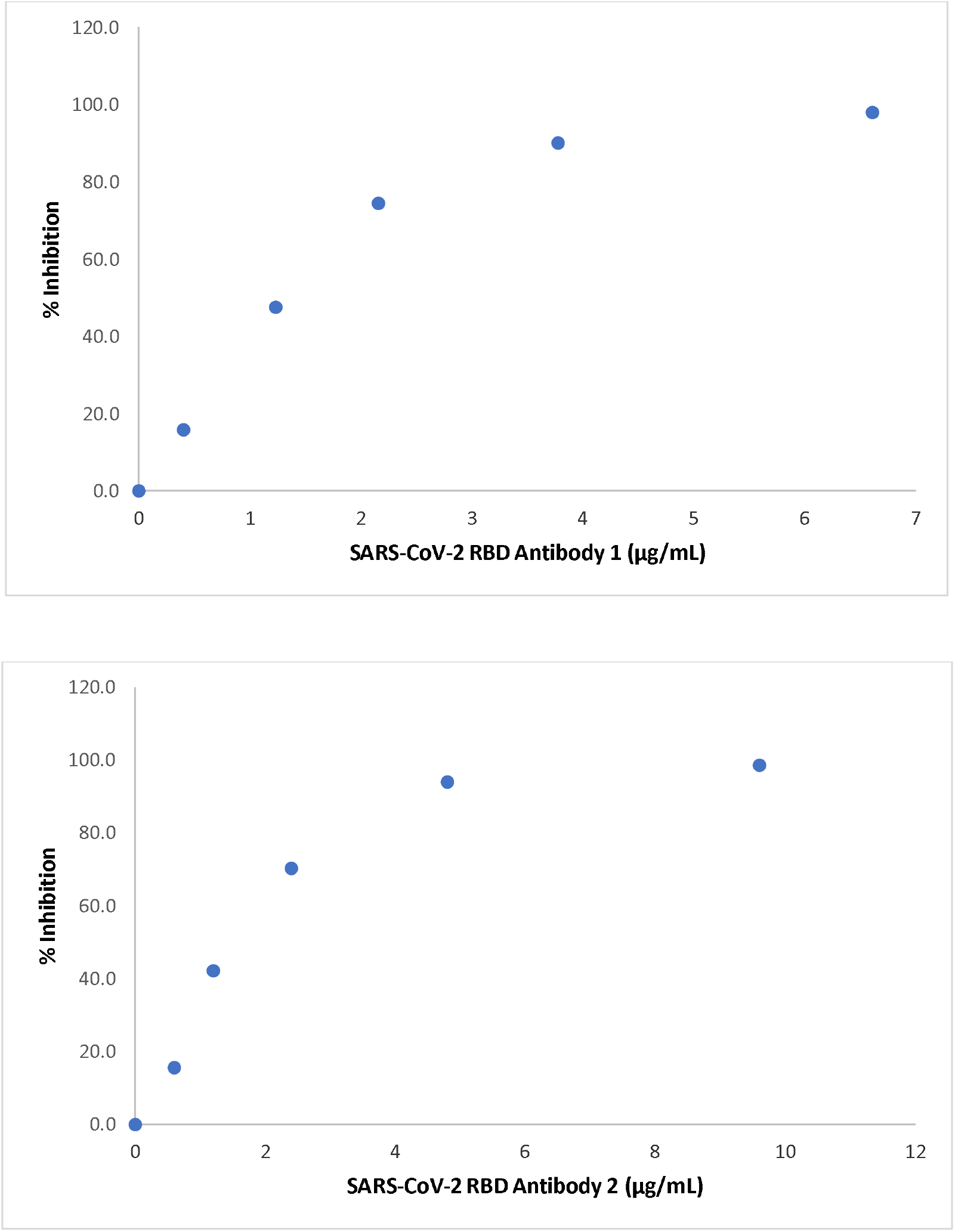
Validation of SARS-CoV-2 Biochip pVNT in a dose response manner. Inhibition of SARS-CoV-2 RBD and ACE2 interaction using specified concentrations of SARS-CoV-2 recombinant neutralising antibodies

### Interrogation of SARS-CoV-2 Biochip pVNT functionality employing longitudinal samples

Longitudinal serum samples were collected at timepoints ranging from 0 days to 15 days post SARS-CoV-2 PCR confirmation and RBD IgG levels were measured using the SARS-CoV-2 IgG (RBD & NP) Array whereas RBD neutralising antibody levels were measured using the SARS-CoV-2 Biochip pVNT (Table 2). Donors 1-3 transitioned to RBD IgG positivity at day 9, day 12 and day 5 post PCR confirmation of SARS-CoV-2 infection respectively. Interestingly, two out of 3 donors transitioned to neutralising antibody positivity at later timepoints when compared to RBD IgG positivity. Donor 2 transitioned at day 15 whereas donor 3 transitioned at day 7.

**Table 2:** Longitudinal assessment of SARS-CoV-2 neutralising antibody levels measured using Biochip SARS-CoV-2 pVNT. Sample collected from SARS-CoV-2 donors confirmed positive by RT-PCR. Results compared to Spike RBD IgG levels.

|  | Day post PCR confirmation | Sample ID | RBD IgG |  | pVNT |  |
| --- | --- | --- | --- | --- | --- | --- |
|  |  |  | SP % | Classification | % inhibition | Classification |
| **Donor 1** | 0 | Sample 1 | 0.0 | Negative | 5.2 | Negative |
|  | 3 | Sample 2 | 0.0 | Negative | -1.0 | Negative |
|  | 6 | Sample 3 | 43.3 | Negative | 4.4 | Negative |
|  | 9 | Sample 4 | 1043.5 | **Positive** | 52.7 | **Positive** |
|  | 12 | Sample 5 | 3259.7 | **Positive** | 88.3 | **Positive** |
|  | 15 | Sample 6 | 3981.0 | **Positive** | 89.2 | **Positive** |
| **Donor 2** | 3 | Sample 1 | 3.4 | Negative | 4.5 | Negative |
|  | 7 | Sample 2 | 6.7 | Negative | 5.5 | Negative |
|  | 8 | Sample 3 | 4.2 | Negative | 11.1 | Negative |
|  | 10 | Sample 4 | 15.0 | Negative | 13.7 | Negative |
|  | 12 | Sample 5 | 307.1 | **Positive** | 27.4 | Negative |
|  | 15 | Sample 6 | 984.8 | **Positive** | 62.1 | **Positive** |
| **Donor 3** | 4 | Sample 1 | 37.7 | Negative | 14.7 | Negative |
|  | 5 | Sample 2 | 72.3 | **Positive** | 16.4 | Negative |
|  | 6 | Sample 3 | 344.0 | **Positive** | 48.0 | Negative |
|  | 7 | Sample 4 | 1093.3 | **Positive** | 59.5 | **Positive** |
|  | 9 | Sample 5 | 2258.3 | **Positive** | 89.0 | **Positive** |
|  | 11 | Sample 6 | 3472.0 | **Positive** | 96.6 | **Positive** |

### Dilutional linearity assessment of the SARS-CoV-2 Biochip pVNT

The newly developed SARS-CoV-2 Biochip pVNT adopts a 1:25 in well sample dilution. Four samples seropositive for SARS-CoV-2 RBD IgG were applied to the SARS-CoV-2 Biochip at dilutions ranging from 1:25 to 1:3200 (Figure 3). All four samples returned a positive result for the presence of SARS-CoV-2 neutralising antibodies when tested at a 1:25 dilution and exhibited a stepwise reduction in % Inhibition as sample input was decreased with increasing sample dilutions. Samples 1-4 transitioned to SARS-CoV-2 antibody negativity at sample dilutions of 1:400, 1:800, 1:800 and 1:800 respectively. These four samples had a microneutralisation titre of 1:261, 1:350, 1:524 and 1:604 respectively.

**Figure 3:**
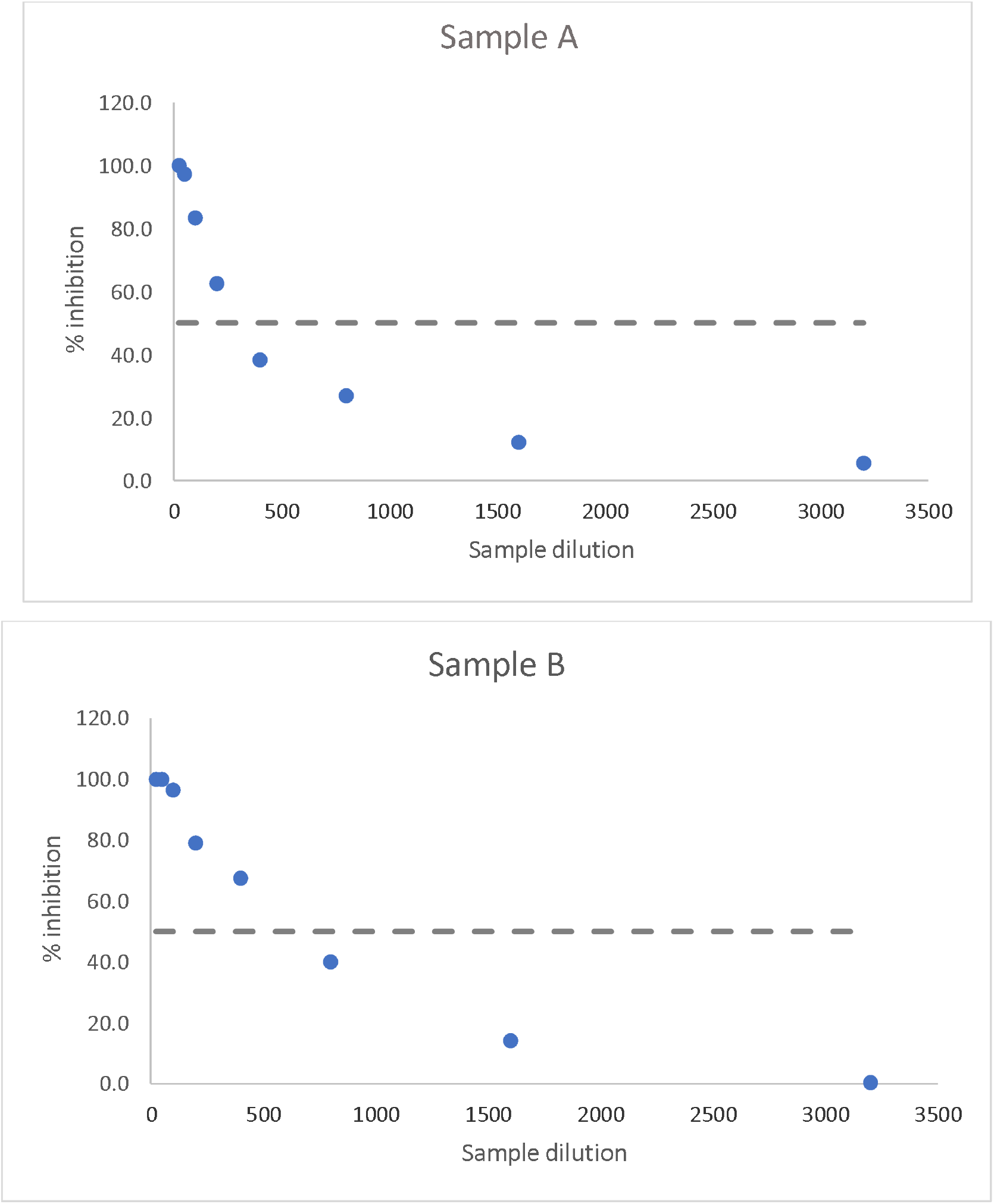
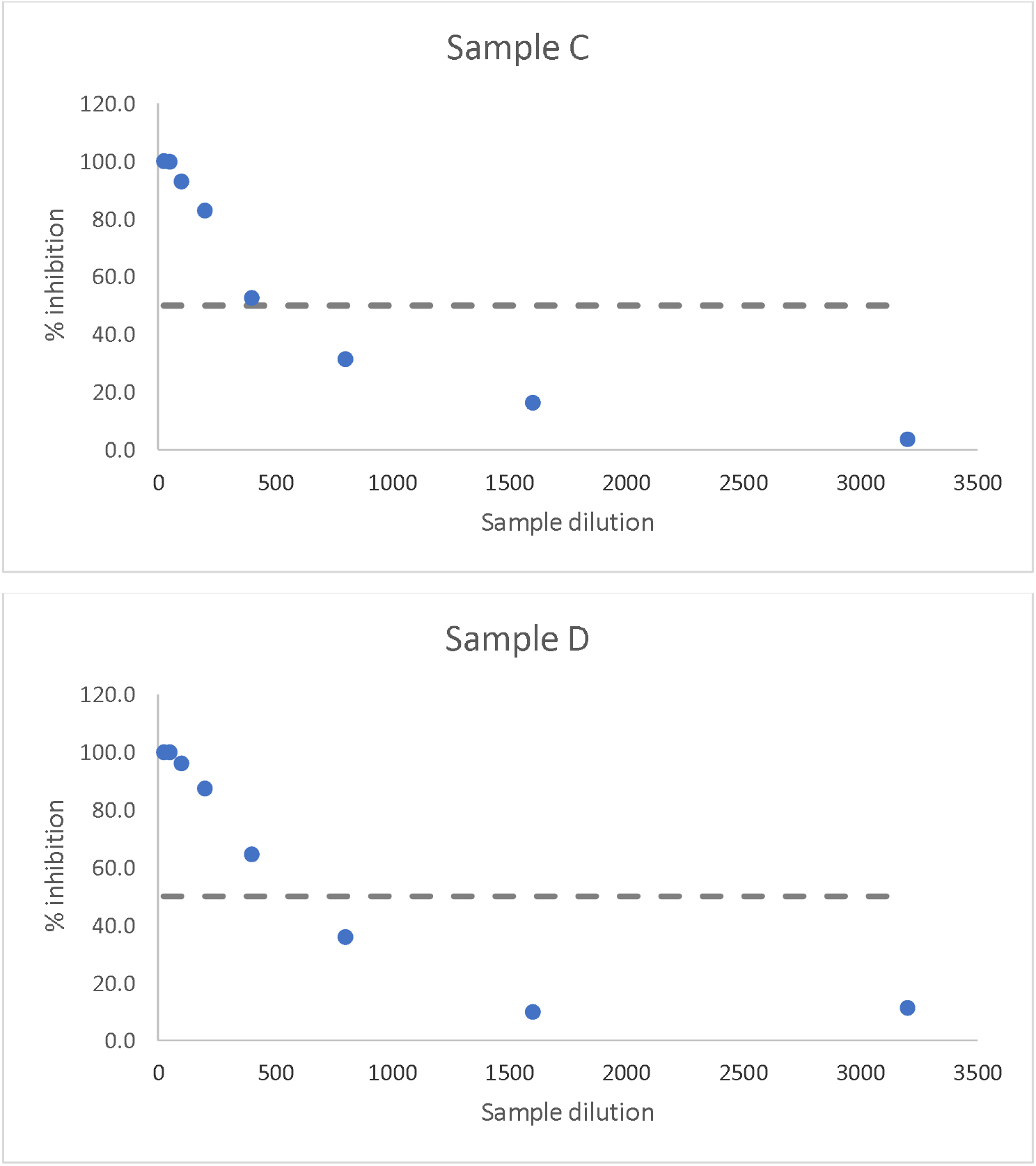
Inhibition of SARS-CoV-2 RBD ACE2 interaction by samples (A-D) collected from patients confirmed neutralising antibody seropositive using the SARS-CoV-2 microneuralisation test. A reduction in % inhibition is observed for all patients using the pVNT as sample dilution increases (starting dilution 1:25). Dotted line indicates SARS-CoV-2 Biochip pVNT cut-off at 50% inhibition.

### Assessment of SARS-CoV-2 Biochip pVNT analytical specificity

The analytical specificity of the SARS-CoV-2 Biochip pVNT was assessed using 169 potentially cross-reacting samples (Table 3). This included samples which were seropositive against all four endemic coronaviruses and 14 additional non-coronavirus pathogens. Excellent analytical specificity was demonstrated in that all 169 samples returned a negative classification for SARS-CoV-2 neutralising antibodies using the SARS-CoV-2 Biochip pVNT (100% analytical specificity).

**Table 3:** Specificity of pVNT against other human coronaviruses and non-coronavirus pathogens

| Category | n | Negative | Positive |
| --- | --- | --- | --- |
| Human coronavirus HKU1 | 9 | 9 | 0 |
| Human coronavirus OC43 | 10 | 10 | 0 |
| Human coronavirus 229E | 10 | 10 | 0 |
| Human coronavirus NL63 | 10 | 10 | 0 |
| Haemophilus influenzae | 18 | 18 | 0 |
| Respiratory syncytial virus (RSV) | 17 | 17 | 0 |
| Influenza A | 3 | 3 | 0 |
| Influenza B | 13 | 13 | 0 |
| Parainfluenza 1-4 | 19 | 19 | 0 |
| Adenovirus | 7 | 7 | 0 |
| Enterovirus | 4 | 4 | 0 |
| *Mycoplasma pneumoniae* | 16 | 16 | 0 |
| Legionella | 5 | 5 | 0 |
| *Chlamydia pneumoniae* | 7 | 7 | 0 |
| Cytomegalovirus | 5 | 5 | 0 |
| Epstein Barr Virus | 6 | 6 | 0 |
| Hepatitis B | 2 | 2 | 0 |
| *Bordetella pertussis* | 8 | 8 | 0 |
| **Total** | **169** | **169** | **0** |

### Correlation of SARS-CoV-2 Biochip pVNT with SARS-CoV-2 microneutralisation assay

The clinical specificity and sensitivity of the SARS-CoV-2 Biochip pVNT was evaluated in a cohort of 110 clinical samples classified as neutralising antibody negative (n=60) or positive (n=50) using the SARS-CoV-2 microneutralisation assay (Figure 4). The SARS-CoV-2 Biochip pVNT demonstrated a clinical specificity of 100% and a clinical sensitivity of 98% when compared to the SARS-CoV-2 microneutralisation assay. Therefore, classification agreement was observed in 109/110 samples with one sample not agreeing between the two methodologies. The discordant sample was classified as neutralising antibody seropositive by the microneutralisation assay with an antibody titre of 1:33. This sample was reported neutralising antibody seronegative using the SARS-CoV-2 Biochip pVNT (47.0% inhibition). An overall correlation coefficient r=0.9355 was obtained when comparing sample classification using both methodologies demonstrating highly comparable performance between both testing approaches.

**Figure 4:**
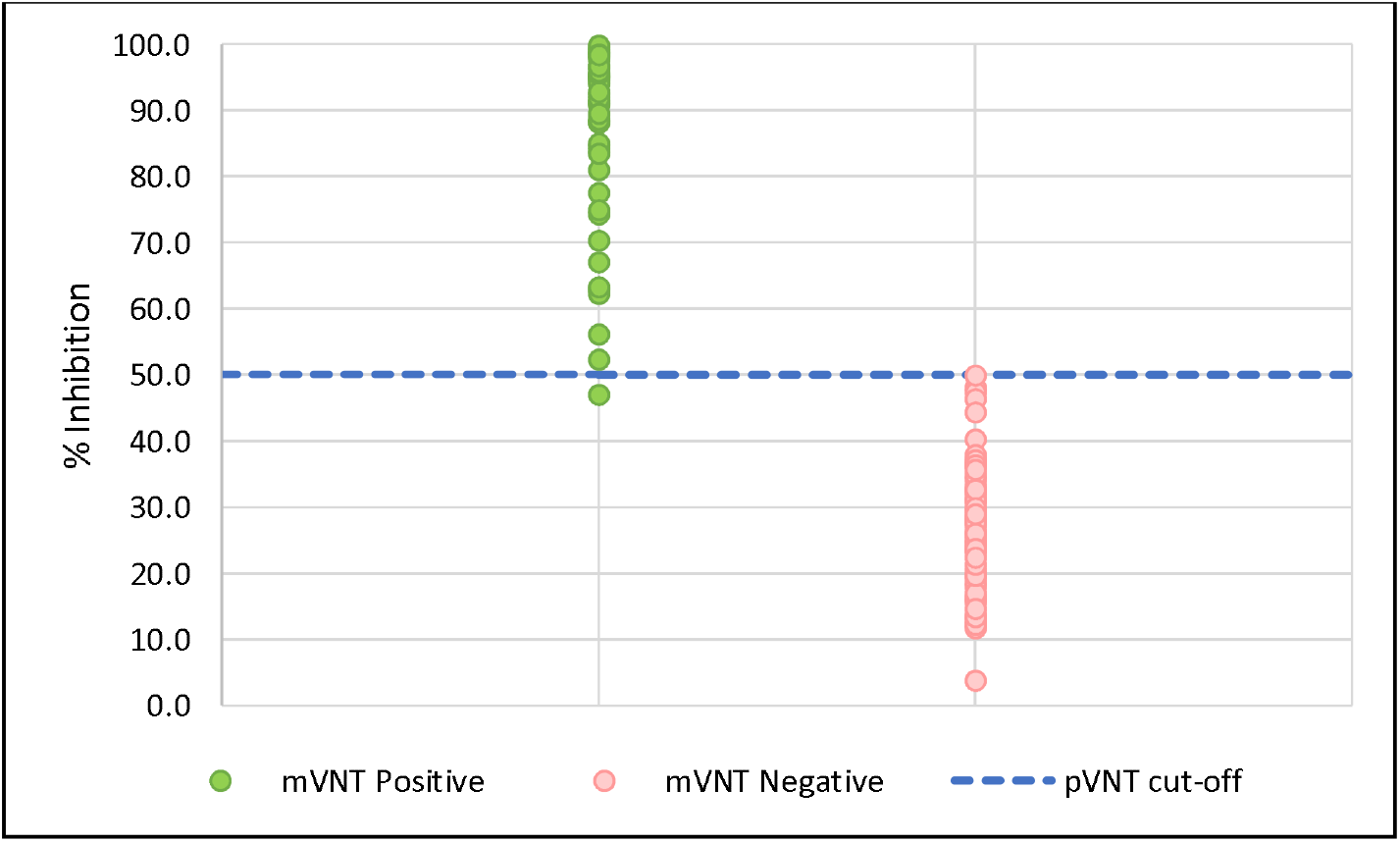
Testing of 110 SARS-CoV-2 microneutralisation assay (mVNT) confirmed seropositive or seronegative samples on the SARS-CoV-2 Biochip pVNT. Dotted line indicates SARS-CoV-2 Biochip pVNT cut-off at 50% inhibition.

### Proof of principle demonstration of species independent applicability of the SARS-CoV-2 Biochip pVNT

Antisera collected from sheep immunized with either SARS-CoV-2 Nucleocapsid protein or SARS-CoV-2 Spike RBD were utilized to investigate potential species independent applicability of the SARS-CoV-2 Biochip pVNT. Antisera was collected at timepoints ranging from day 0 to day 52 post immunization and applied to the SARS-CoV-2 Biochip pVNT (Table 4). Interestingly, antisera from sheep immunized with RBD at day 52 exhibited 100.0% inhibition of the RBD/ACE2 interaction. Antisera from earlier timepoints appear negative for neutralising antibodies exhibiting 28.8% inhibition at day 6 for example. The specificity of this experimental approach was demonstrated by the observation that antisera from sheep immunized with Nucleocapsid protein were consistently negative for neutralising antibodies at all timepoints evaluated (0-52 days). This data strongly indicates the species independent applicability of the newly developed SARS-CoV-2 Biochip pVNT.

**Table 4:**
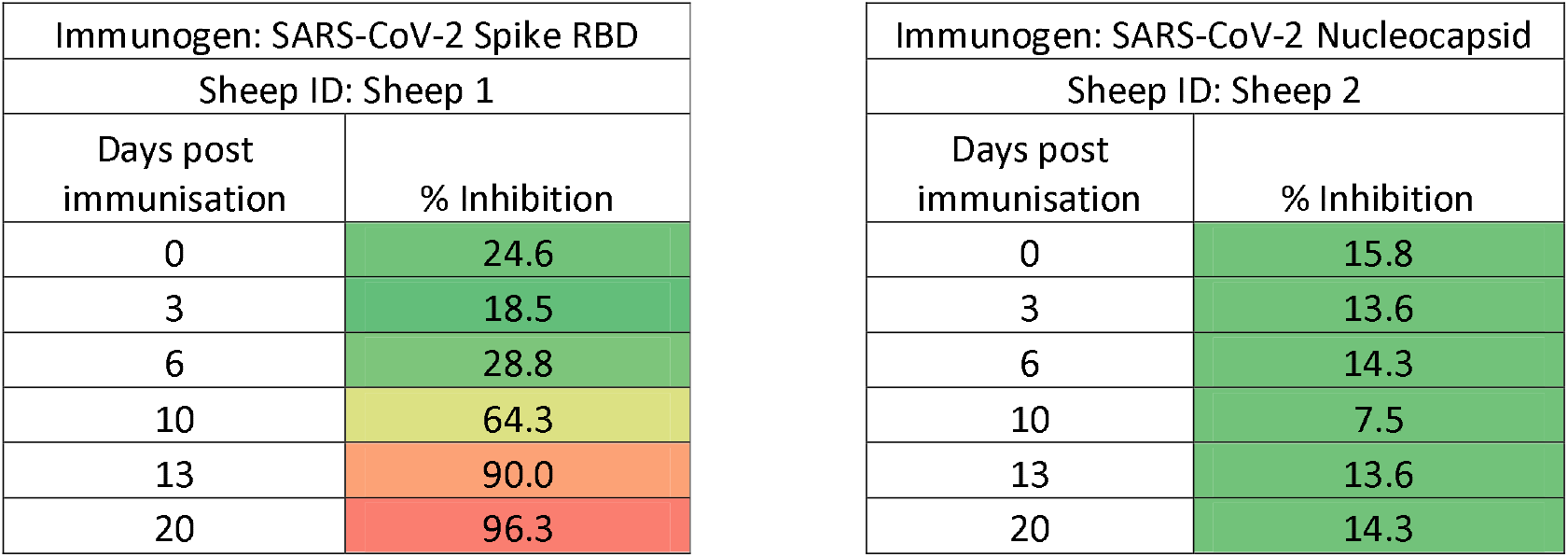
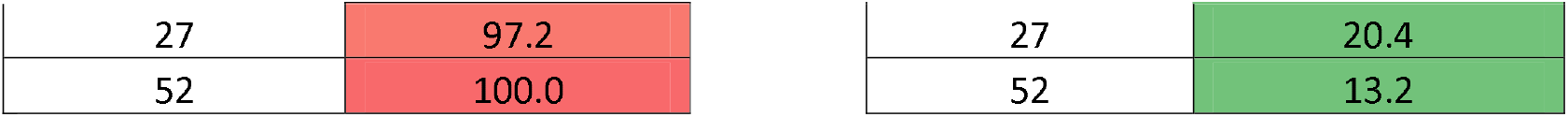
Species independent SAR-CoV-2 neutralising antibody detection using SARS-CoV-2 Biochip pVNT

## Discussion

Randox has developed a rapid Biochip pVNT for the detection of neutralising antibodies against SARS-CoV-2 amendable to high throughput testing in a routine clinical laboratory setting. This contrasts classical virus neutralisation antibody detection methods whereby barriers to rapid and high throughput testing include the requirement for live virus utilisation, highly trained laboratory personnel and BSL3 facilities^10^. The SARS-CoV-2 Biochip pVNT can deliver results within 1.5 hours compared to the 2-4 days required for the virus neutralisation test. Given that antibodies targeting SARS-CoV-2 Spike RBD account for >90% neutralising antibody activity in convalescent sera^11-12^ and that neutralising antibodies largely correlate with protection^13-14^, the widespread availability of rapid and amenable neutralising antibody tests is necessary to accurately ascertain measurements of global herd immunity post natural infection and vaccination. This is fundamental in accurately quantifying risk of infection and onwards virus transmission. Current sero-surveillance is primarily carried out using SARS-CoV-2 serology tests which can be either IgG specific or measure total Ig (IgG, IgA, IgM) levels. This strategy, while worthwhile, has some limitations as such approaches cannot differentiate neutralising from non-neutralising antibodies and results obtained do not equate to protective immunity. Therefore, the SARS-CoV-2 Biochip pVNT test described in this study can positively impact current testing deficiencies in this area.

ACE2 represents the primary receptor for viral entry into human cells and multiple studies have demonstrated that it is the RBD within the Spike protein of SARS-CoV-2 that interacts with ACE2 to precipitate viral entry^15^. During this study, we explored a pVNT format whereby ACE2 was immobilised on the Biochip surface and an alternative pVNT assay format whereby Spike RBD was immobilised on the Biochip surface. Good agreement in the detection of neutralising antibodies against SARS-CoV-2 was observed as demonstrated using recombinant neutralising antibodies and SARS-CoV-2 seropositive clinical samples. (Table 1A, 1B). The putative assay format whereby ACE2 is immobilized on Biochip surface requires a sample pre-incubation step where samples must be incubated off board with HRP labelled RBD prior to addition to ACE2 immobilized on the Biochip surface. This pre-incubation step is cumbersome with respect to manual immunoassay running, potentially increases the possibility of error due to increased immunoassay complexity and poses additional challenges with respect to achieving maximum throughput on automated immunoassay platforms. The alternative format (Spike RBD immobilised on Biochip surface) now utilised in the SARS-CoV-2 Biochip pVNT eliminates the requirement for an offboard sample pre-incubation step, simplifying manual immunoassay running and provides advantages with respect to achieving maximum throughput on automated immunoassay platforms, an important consideration with respect to the current status of the COVID-19 pandemic.

In this study an excellent correlation between the SARS-CoV-2 Biochip pVNT and the SARS-CoV-2 microneutralisation test was observed. All 60 SARS-CoV-2 seronegative samples included in the correlation evaluation along with all 169 potential cross reactant samples were classified correctly using the SARS-CoV-2 Biochip pVNT. Forty nine of the 50 samples classified as seropositive using the SARS-CoV-2 microneutralisation assay were also detected using the SARS-CoV-2 Biochip pVNT. The single discordant sample which was a weak positive using the SARS-CoV-2 microneutralisation assay reported 47% inhibition using the SARS-CoV-2 Biochip pVNT thus just below the pVNT assay threshold (50%). The serial dilution of four samples (Figure 3) with high neutralising antibody levels reported an end-point dilution using the SARS-CoV-2 Biochip comparable with that obtained using the SARS-CoV-2 microneutralisation test. This demonstrates the potential of the SARS-CoV-2 Biochip pVNT to monitor longitudinal neutralising antibody responses in a semi-quantitative fashion. It was also interesting to note that in clinical samples RBD IgG levels did not directly correlate with neutralising antibody activity. Thus, corroborating the hypothesis that measurement of RBD IgG levels does not necessarily infer the presence of protective/neutralising antibodies to SARS-CoV-2. These results further exemplifying the utility of the newly developed SARS-CoV-2 Biochip pVNT as a robust and reliable method for the accurate measurement of neutralising antibodies against SARS-CoV-2.

Another significant advantage of the SARS-CoV-2 Biochip pVNT is its ability to detect neutralising antibodies in a species independent manner. The virus is presumed of zoonotic origin^16^ and there are growing concerns regarding SARS-CoV-2 animal reservoirs and their potential to precipitate future outbreaks. It has been demonstrated that diverse ACE2 orthologs from different animal species can support SARS-CoV-2 infection^17-22^ and like humans, SARS-CoV-2 neutralsing antibodies can protect against infection^23^. In this study, the SARS-CoV-2 Biochip pVNT demonstrates the ability to detect a SARS-COV-2 neutralsing antibody response in sheep with a significant increase in neutralising antibody response observed by day 10 post SARS-CoV-2 Spike RBD immunisation whereas no neutralising antibody response was observed following Nucleocapsid protein immunisation. Improved diagnostic focus supportive of the “One Health” approach to infectious disease along with increased surveillance is required to fully understand the underlying mechanisms involved in cross-species transmission and to control the significant risk posed by animal reservoirs in the spread of SARS-CoV-2.

SARS-CoV-2 Spike RBD is a major target in vaccine development. The measurement of neutralising antibodies post vaccination provides supportive evidence in assessing vaccine efficacy and also assists with the evaluation of future vaccine candidates. For a SARS-CoV-2 vaccine to be effective it must stimulate sufficient antibody production to block the interaction between Spike RBD and ACE2. Where a mismatch between the vaccine and circulating virus strain occurs this can have a significant impact on vaccine elicited neutralisation^24^. The SARS-CoV-2 Biochip pVNT provides an accessible and effective means of monitoring response to vaccination and for the screening and identification of effective monoclonal antibody therapies. This assay is highly adaptable and can be modified to include Spike RBD variants representative of displaced or evolving dominant SARS-CoV-2 virus lineages as required.

To conclude, the dynamics of the neutralising antibody response in patients who have recovered from or have been vaccinated against SARS-CoV-2 can vary greatly and estimates of protection and immune longevity can only accurately be determined at an individual level. The availability of a rapid, robust and reliable high throughput SARS-CoV-2 Biochip pVNT can assist in providing accurate measurements of immunity at a population level and inform public health decisions moving forward with regards appropriate vaccination regimes and the necessity to update vaccine strains. The SARS-CoV-2 Biochip pVNT can also assist with expanding our current limited knowledge on the understanding of how this virus interacts with the immune response and can migrate within various animal populations. All of the above are critical aspects in the management of this evolving virus and in the management of future SARS-CoV-2 virus variants.

## Data Availability

The data that support the findings of this study are available from the corresponding author, CR, upon reasonable request.

